# Mental health of Adolescents in the Pandemic: Long-COVID19 or Long-Pandemic Syndrome?

**DOI:** 10.1101/2021.05.11.21257037

**Authors:** Judith Blankenburg, Magdalena K. Wekenborg, Jörg Reichert, Carolin Kirsten, Elisabeth Kahre, Luise Haag, Leonie Schumm, Paula Czyborra, Reinhard Berner, Jakob P. Armann

## Abstract

**Backround:** Post-COVID19 complications such as pediatric inflammatory multisystem syndrome (PIMS) and Long-COVID19 move increasingly into focus, potentially causing more harm in this age group than the acute infection. To better understand the symptoms of long-COVID19 in adolescents and to distinguish infection-associated symptoms from pandemic-associated symptoms, we conducted a Long-COVID19 survey, comparing responses from seropositive and seronegative adolescents. To our knowledge, data of Long-COVID19 surveys with seronegative control groups have not been published yet.

**Methods:** Since May 2020 students grade 8-12 in fourteen secondary schools in Eastern Saxony were enrolled in the SchoolCovid19 study. Seroprevalence was assessed via serial SARS-CoV-2 antibody testing in all participants. Furthermore, during the March/April 2021 study visit all participants were asked to complete a 12 question Long-COVID19 survey regarding the occurrence and frequency of difficulties concentrating, memory loss, listlessness, headache, abdominal pain, myalgia/ arthralgia, fatigue, insomnia and mood (sadness, anger, happiness and tenseness).

**Findings:** 1560 students with a median age of 15 years participated in this study. 1365 (88%) were seronegative, 188 (12%) were seropositive. Each symptom was present in at least 35% of the students within the last seven days before the survey. However, there was no statistical difference comparing the reported symptoms between seropositive students and seronegative students. Whether the infection was known or unknown to the participant did not influence the prevalence of symptoms.

**Interpretation:** The lack of differences comparing the reported symptoms between seropositive and seronegative students suggests that Long-COVID19 might be less common than previously thought and emphasizing the impact of pandemic-associated symptoms regarding the well-being and mental health of young adolescents.

**Funding:** This study was supported by a grant by the Federal State of Saxony. M.K.W. was supported by the Else Kröner-Fresenius Center for Digital Health (EKFZ), TU Dresden, Germany.

## Introduction

Since the identification of the severe acute respiratory syndrome coronavirus 2 (SARS-CoV-2) as the cause of COVID-19 ^1^ in December 2019 and the beginning of the SARS-CoV-2 pandemic in Germany in March 2020 nearly 490.000 cases in children and adolescents have been reported by the Robert-Koch-Institute (RKI)^2^. In contrast to adults, children and adolescents usually have mild disease courses with a low rate of hospitalization^3–5^. Therefore, post-COVID19 complications such as pediatric inflammatory multisystem syndrome (PIMS) and Long-COVID19 -with persisting symptoms 4 – 12 weeks and more than 12 weeks after an acute SARS-CoV-2infection^6^ move into focus, potentially causing more harm in this age group than the acute infection.

While multiple studies and registers have provided reliable data on epidemiology, clinical presentation, disease course and treatment options on PIMS^7–9^ to date no comparable data exists for Long-COVID19 in children and adolescents. A cross-sectional study from Italy^10^ in 123 children diagnosed with a SARS-CoV-2 infection found that more than 50% of participants had at least one persisting symptom 120 days after their infection, with Insomnia, pain, fatigue, and concentration difficulties being the most commonly reported ones. An April 2021 Office for National Statistics (ONS) report from the UK^11^ similarly provided data that symptoms in children persisted at least 12 weeks after their SARS-CoV-2 infection.

These numbers are concerning and require attention; however, currently they merely show a temporal connection and not a causal relationship. In order to better understand the epidemiology and clinical manifestations of Long-COVID19 in children and adolescents and differentiate infection-associated symptoms from pandemic-associated symptoms, we conducted a Long-COVID19 survey in more than 1500 students participating in the SchoolCoviDD19 study in March and April 2021.

## Methods

### Study Design

Since May 2020 students grade 8-12 in fourteen secondary schools in Eastern Saxony are enrolled in the SchoolCoviDD19study. Two of these 14 schools are vocational schools. Seroprevalence is assessed via serial SARS-CoV-2 antibody assessment in all participants. During the March/April 2021 study visit all participants were asked to complete a Long-COVID19 survey. Vaccinated Students (n=7) were excluded from the analysis.

### Survey Details

The Survey included besides sociodemographic variables (i.e., age, sex) twelve questions on the occurrence and frequency of relevant neurocognitive, pain and mood symptoms, namely difficulties concentrating, memory loss, listlessness, headache, abdominal pain, myalgia/arthralgia, fatigue, insomnia and mood (sadness, anger, happiness and tenseness) within the last seven days before the survey.

The questions were taken from the Symptom Checklist-90-R (SCL-90-R)^12^, the Somatic Symptom Scale (SSS-8)^13^ and a questionnaire about stress and stress management in children and adolescents (SS KJ 3-8 R)^14^. All questionnaires are validated in adolescents.

Answers were coded on a categorical scale – “never”, “once”, “multiple times” for insomnia and all mood questions; “not at all”, “a little bit”, “quite”, “severe” and “very severe” for the remaining questions.

In addition, a self-generated item was used to assess the overall level of mental distress on a scale from 0 (“not at all”) to 10 (“total”).

### Laboratory Analysis

We assessed anti-SARS-CoV-2 IgG antibodies in all samples using a commercially available chemiluminescence immunoassay (CLIA) technology for the quantitative determination of anti-S1 and anti-S2 specific IgG antibodies to SARS-CoV-2 (Diasorin LIAISON® SARS-CoV-2 S1/S2 IgG Assay). Antibody levels > 15·0 AU/ml were considered positive and levels between 12·0 and 15·0 AU/ml were considered equivocal.

All samples with a positive or equivocal LIAISON® test result, as well as all samples from participants with a reported personal or household history of a SARS-CoV-2 infection, were re-tested with two additional serological tests. These were a chemiluminescent microparticle immunoassay (CMIA) intended for the qualitative detection of IgG antibodies to the nucleocapsid protein of SARS-CoV-2 (Abbott Diagnostics® ARCHITECT SARS-CoV-2 IgG) (an index (S/C) of < 1·4 was considered negative whereas one >/= 1·4 was considered positive) and an ELISA detecting IgG against the S1 domain of the SARS-CoV-2 spike protein (Euroimmun® Anti-SARS-CoV-2 ELISA) (a ratio < 0·8 was considered negative, 0·8–1·1 equivocal, > 1·1 positive).

Participants whose positive or equivocal LIAISON® test result could be confirmed by a positive test result in at least one additional serological test were considered seropositive. Participants with a negative LIAISON® test result, but positive results in both additional serological tests were also considered positive.

### Statistical Analysis

Results for continuous variables are presented as medians with interquartile ranges (IQR) and categorical variables as numbers with percentages, unless stated otherwise.

Fisher’s exact test was used to determine categorical variables for the statistical analysis. Thereby, the answers to the items assessing neurocognitive, pain and mood symptoms, were dummy-coded, enabling a comparison of the answer category “none” (coded 0) against the answer categories “a little bit”/ “quite”/ “severe”/ “very severe” and “once”/ “multiple times” (coded 1), respectively.

Furthermore, data distributions of the neurocognitive, pain and mood symptoms were tested for normality using the Kolmogorov-Smirnov (K-S) test. Data with distributions significantly different (p< 0·05) from normal were either transformed to ranks to allow parametric statistics or analyzed using non-parametric statistics.

In order to examine associations between sociodemographic variables (i.e., age, sex) and the neurocognitive and pain symptoms, bivariate correlation analyses were conducted.

In a second step, partial correlation analyses were conducted between serostatus and the neurocognitive and pain symptoms, adjusting for age and sex.

Analyses were performed using IBM SPSS 27.0 or Microsoft Excel 2010. All statistical tests were conducted with α < 0.05.

### Approval

The SchoolCoviDD19 study was approved by the Ethics Committee of the Technische Universität (TU) Dresden (BO-EK-156042020) and has been assigned clinical trial number DRKS00022455.

## Results

1560 students with a median age of 15 years participated at the study visit in March/April 2021 and had their serostatus analyzed. Seven already vaccinated students were excluded from the analysis, 1365 (88%) were seronegative and 188 (12%) were seropositive. Median age, sex and household size did not differ significantly between seropositive and seronegative participants (*table 1*).

**Table 1).**
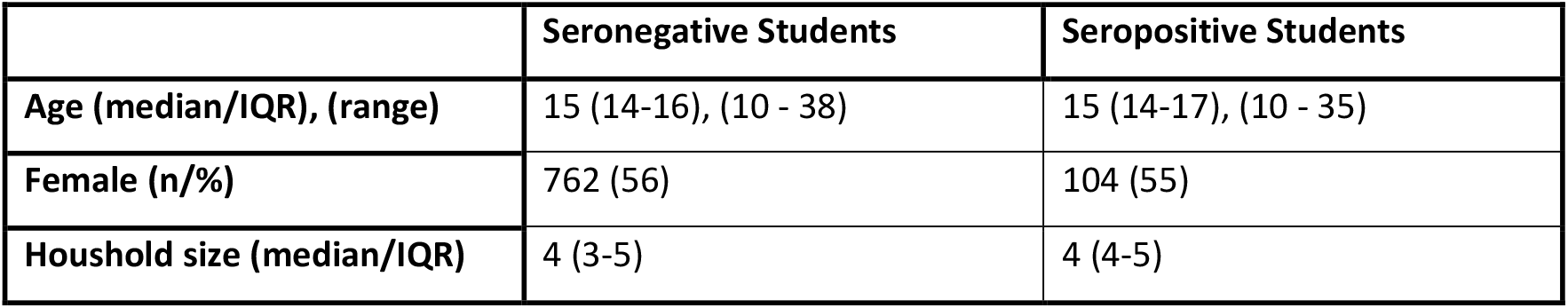
Demographic data and serostatus of the participating students at study visit in March/ April 2021; IQR – Interquartile range

|  | Seronegative Students | Seropositive Students |
| --- | --- | --- |
| Age (median/IQR), (range) | 15 (14-16), (10 - 38) | 15 (14-17), (10 - 35) |
| Female (n/%) | 762 (56) | 104 (55) |
| Houshold size (median/IQR) | 4 (3-5) | 4 (4-5) |

Long-COVID19 survey was answered by at least 1504 (96·8%) of the participants. Each symptom, regardless of the expression, was present in at least 35% of the students within the last seven days before the survey, most commonly happiness (98·7%) followed by tenseness (86·4%), listlessness (80·7%) and difficulties concentrating (79·3%). Myalgia/arthralgia (35·6%) and fatigue (37·8%) were reported least commonly. (see *table 2* for full results).

**Table 2).**
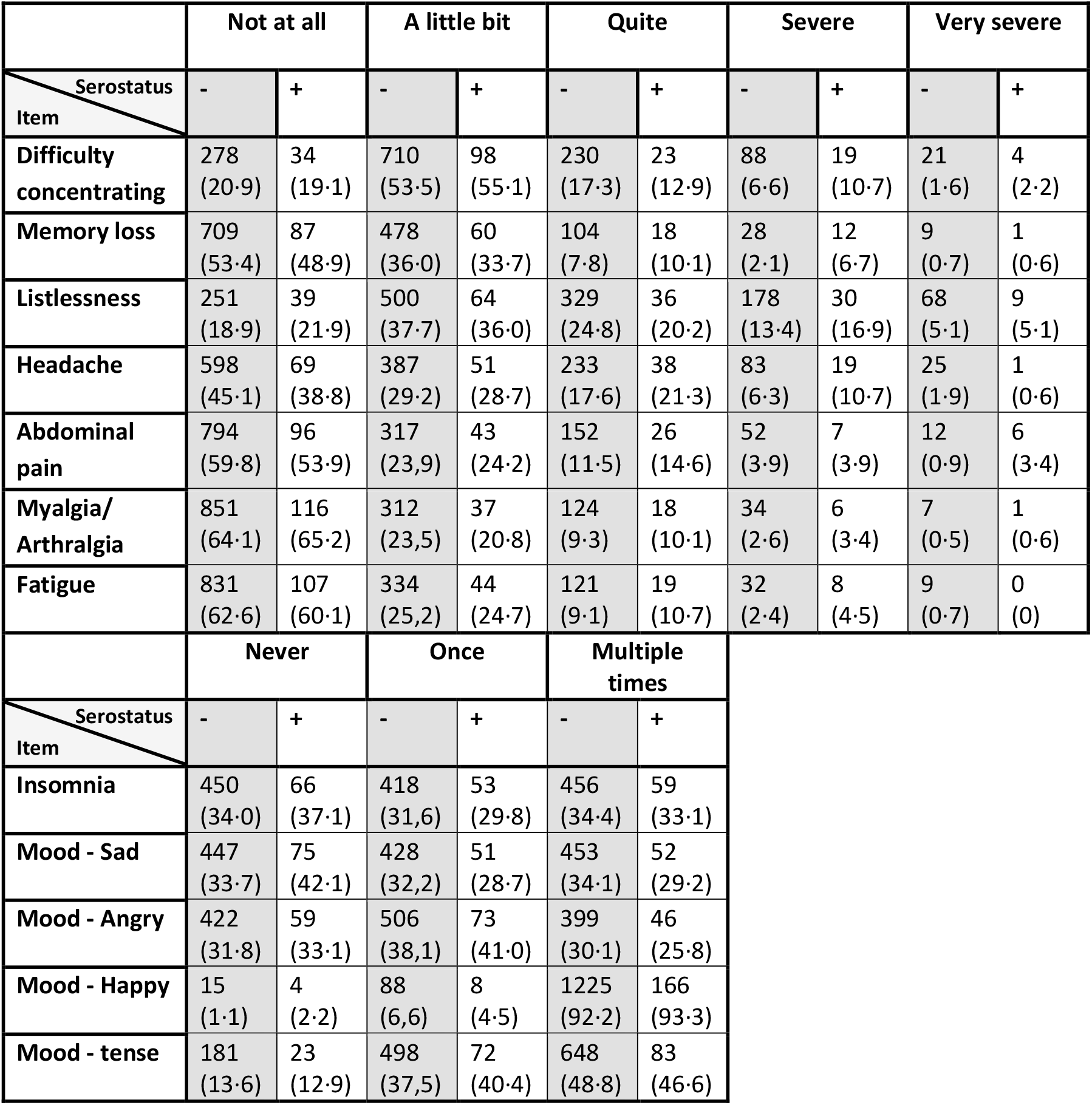
All answers of seronegative (-) and seropositive (+) participants to the Long-COVID19-survey

Fisher’s exact test did not reveal any significant differences between seropositive and seronegative students regarding the prevalence of any of the neurocognitive and pain symptoms reported (*figure 1*).

**Figure 1).**
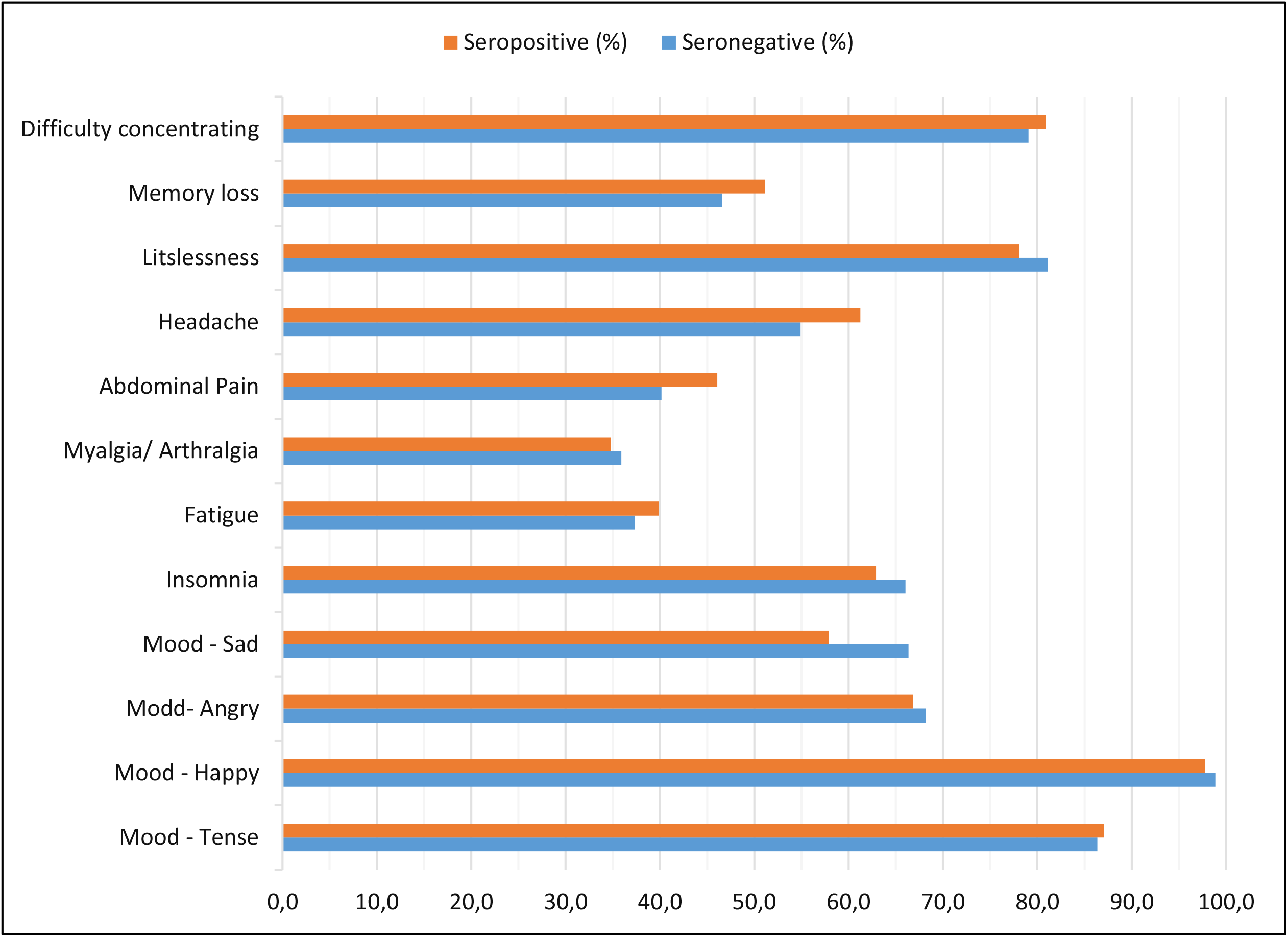
Prevalence of neurocognitive, pain and mood symptoms in seronegative and seropositive study participants *(Fisher’s exact test:* n = 1553, * significant at level 0·05)

To avoid underestimation of seropositive individuals due to our relatively strict definitions of seropositivity, we also analyzed the data if only the LIAISON® test result was taken into consideration for the decision on seropositivity. This resulted in 204 (13%) LIAISON®-seropositive and 1342 (86%) -seronegative students. There was no statistical difference (Fisher’s exact test) in the occurrence of any neurocognitive, pain or mood symptoms between these LIAISON®-seropositive and – seronegative students either (see S*upplementals - figure 1*).

Spearman correlation analyses revealed that age was positive correlate with nearly all neurocognitive and pain symptoms, except for insomnia, sad mood and angry mood (*table 3*). In addition, females reported a consistently higher prevalence of neurocognitive and pain symptoms compared to men, except for Myalgia Arthralgia where there was no significant association with sex.

**Table 3).** *Spearman-Rho bivariate correlations between age, sex, and the reported neurocognitive, pain and mood symptoms* (n = 1553, * significant at level 0·05, ** significant at level 0·01 (one-tailed test))

| Variable | (1) | (2) | (3) | (4) | (5) | (6) | (7) | (8) | (9) | (10) | (11) | (12) | (13) | (14) |
| --- | --- | --- | --- | --- | --- | --- | --- | --- | --- | --- | --- | --- | --- | --- |
| (1) Age |  |  |  |  |  |  |  |  |  |  |  |  |  |  |
| (2) sex | -.05 |  |  |  |  |  |  |  |  |  |  |  |  |  |
| (3) Distress | .23** | .25** |  |  |  |  |  |  |  |  |  |  |  |  |
| (4) Concentration | .08** | .14** | .35** |  |  |  |  |  |  |  |  |  |  |  |
| (5) Memory | .10** | .07** | .24** | .47** |  |  |  |  |  |  |  |  |  |  |
| (6) Listlessness | .17** | .08** | .36** | .52** | .32** |  |  |  |  |  |  |  |  |  |
| (7) Headache | .10** | .27** | .30** | .30** | .23** | .27** |  |  |  |  |  |  |  |  |
| (8) Abdominal Pain | .05* | .21** | .26** | .21** | .20** | .23** | .28** |  |  |  |  |  |  |  |
| (9) Myalgia Arthralgia | -.07* | .05 | .15** | .24** | .21** | .20** | .21** | .26** |  |  |  |  |  |  |
| (10) Fatigue | .11** | .10** | .26** | .32** | .25** | .29** | .28** | .27** | .35** |  |  |  |  |  |
| (11) Insomina | .04 | .20** | .30** | .28** | .23** | .22** | .29** | .18** | .18** | .24** |  |  |  |  |
| (12) Sad | .04 | .38** | .48** | .35** | .27** | .33** | .30** | .28** | .18** | .27** | .37** |  |  |  |
| (13) Angry | -.04 | .12** | .24** | .23** | .20** | .22** | .17** | .18** | .14** | .16** | .23** | .33** |  |  |
| (14) Happy | -.05* | .06* | -.11* | -.17** | -.12** | -.17** | -.06* | -.05* | -.07** | -.11** | -.12** | -.11** | -.06* |  |
| (15) Tense | .14** | .16** | .33** | .29** | .23** | .30** | .20** | .20** | .14** | .23** | .25** | .36** | .28** | -.07** |

Partial correlation analyses, which were performed to test for age and sex independent effects of the analysed serostatus on rank-transformed neurocognitive and pain symptoms revealed differences only with respect to sadness; with being seronegative was associated with an increased prevalence of sadness (*table 4*).

**Table 4).** *Partial correlations between serostatus and neurocognitive, pain and mood symptoms (rank-transformed), controlling for age and sex* (n = 1553, * significant at level 0·05, ** significant at level 0·01 (one-tailed test))

| Variable | (1) | (2) | (3) | (4) | (5) | (6) | (7) | (8) | (9) | (10) | (11) | (12) | (13) |
| --- | --- | --- | --- | --- | --- | --- | --- | --- | --- | --- | --- | --- | --- |
| (1) Serostatus |  |  |  |  |  |  |  |  |  |  |  |  |  |
| (2) Mental Distress | -.01 |  |  |  |  |  |  |  |  |  |  |  |  |
| (3) Concentration | .02 | .33** |  |  |  |  |  |  |  |  |  |  |  |
| (4) Memory | .05 | .22** | .47** |  |  |  |  |  |  |  |  |  |  |
| (5) Listlessness | -.01 | .34** | .51** | .31** |  |  |  |  |  |  |  |  |  |
| (6) Headache | .05 | .24** | .27** | .21** | .24** |  |  |  |  |  |  |  |  |
| (7) Abdominal Pain | .05 | .22** | .18** | .18** | .22** | .23** |  |  |  |  |  |  |  |
| (8) Myalgia Arthralgia | <-.01 | .16** | .23** | .21** | .20** | .20** | .26** |  |  |  |  |  |  |
| (9) Fatigue | .02 | .23** | .31** | .24** | .27** | .25** | .25** | .35** |  |  |  |  |  |
| (10) Insomina | -.02 | .26** | .26** | .22** | .20** | .25** | .14** | .18** | .23** |  |  |  |  |
| (11) Sad | -.06* | .44** | .32** | .25** | .32** | .22** | .22** | .17** | .25** | .32** |  |  |  |
| (12) Angry | -.02 | .22** | .22** | .20** | .21** | .15** | .18** | .13** | .15** | .22** | .31** |  |  |
| (13) Happy | .01 | -.12** | -.17** | -.12** | -.17** | -.07** | -.07* | -.08** | -.11** | -.13** | -.14** | -.07* |  |
| (14) Tense | -.02 | .28** | .27** | .22** | .28** | .16** | .17** | .15** | .21** | .22** | .31** | .27** | -.07** |

104 out 188 seropositive students (55%) had previously been tested positive for SARS-CoV-2 and/ or reported a confirmed SARS-CoV-2 positive household member and were therefore considered as known SARS-CoV2 infections. Compared to those with an unknown infection (84/188 (45%)) Fisher’s exact test did not reveal any significant differences regarding the prevalence of any of the neurocognitive and pain symptoms reported either (*table 5*).

**Table 5).** *Answers of seropositive participants to the Long-COVID19-survey – known vs. previously unknown infection (Fisher’s exact test:* n = 1553, * significant at level 0·05)

|  | None |  | Any |  | p |
| --- | --- | --- | --- | --- | --- |
|  | known infection | unknown infection | known infection | unknown infection |  |
| **Difficulty concentrating** | 22 (22.5) | 12 (15.0) | 76 (77.6) | 68 (85.0) | NS |
| **Memory loss** | 47 (48.0) | 40 (50.0) | 51 (52.0) | 40 (50.0) | NS |
| **Listlessness** | 21 (21.4) | 18 (22.5) | 77 (78.6) | 62 (77.5) | NS |
| **Headache** | 40 (40.8) | 29 (36.3) | 58 (59.2) | 51 (63.8) | NS |
| **Abdominal pain** | 55 (56.1) | 41 (51.2) | 43 (43.9) | 39 (48.8) | NS |
| **Myalgia/Arthralgia** | 64 (65.3) | 52 (65.0) | 34 (34.7) | 28 (35.0) | NS |
| **Fatigue** | 61 (62.2) | 46 (57.5) | 37 (37.8) | 34 (42.5) | NS |
|  | Never |  | At least once |  | p |
|  | known infection | unknown infection | known infection | unknown infection |  |
| **Insomnia** | 35 (35.7) | 31 (38.8) | 63 (64.3) | 49 (61.3%) | NS |
| **Mood - Sad** | 41 (41.8) | 34 (42.5) | 57 (58.2) | 46 (57.58) | NS |
| **Mood - Angry** | 30 (30.6) | 29 (36.3) | 68 (69.4) | 51 (63.8) | NS |
| **Mood – Happy** | 1 (1.0) | 3 (3.8) | 97 (99.0) | 77 (96.3) | NS |
| **Mood - Tense** | 11 (1.22) | 12 (15.0) | 87 (88.8) | 68 (85.0) | NS |

The median score of self-reported mental distress was 4 and did not differ between seropositive and seronegative participants.

## Discussion

The data presented in our study clearly shows a high rate of neurocognitive, pain and mood symptoms in the surveyed group of adolescents, with every item being present in at least one third of the students within the last seven days before responding to the survey. This is consistent with previous studies and surveys on the prevalence of Long-COVID19 symptoms^10^ or psychosomatic symptoms during the SARS-CoV-2 pandemic^15^ in this age group. Furthermore the prevalence is considerably higher compared to pre-pandemic data.^15^

Our study can know provide a control group to SARS-CoV-2 infected adolescents by comparing the responses of seropositive individuals to those of their seronegative peers which has not been published so far.

The differentiation between infection-associated and pandemic-associated symptoms is important because the approach to mediate these symptoms will be different. While strict lock-down measures including school closures will prevent SARS-CoV-2 transmissions in this age group and thereby prevent long-term infection related illnesses, these measures will also further restrict social contact, self-determination, education and development of the affected children and adolescents and thereby amplify pandemic-or lockdown-associated symptoms.

The equal prevalence of neurocognitive, pain and mood symptoms in seronegative and seropositive adolescents in our study does not negate the existence of Long-COVID19 symptoms in general or in the pediatric population. However, it does suggest that they occur less frequently than previously assumed – at least in children and adolescents with only mild to asymptomatic courses of disease – as were investigated by this study.

Furthermore, it confirms the negative effects of lockdown measures on mental health and well-being of children and adolescents^16^. These effects – affecting this whole age group – need to be balanced with the risk of Long-COVID19 in infected individuals. This balancing act will be a difficult task for public health officials and political officials. Nevertheless, it will be a necessary one when aiming to improve mental health in adolescents.

While self-reported symptoms cannot be equated with the diagnosis of an illness, a prevalence of at least 35% for each symptom is a concerning screening result that requires further investigation. In addition, validated, reliable tests are needed to evaluate symptom severity in affected individuals. The fact that self-reported overall mental distress did not differ significantly between seropositive and seronegative individuals does not suggest though that infection-associated symptoms are necessarily more severe than pandemic associated symptoms. The interpretation of the negative correlation of sadness and positive serostatus in the partial correlation analyses is difficult and should be but not the overstated given the fact that the group (none vs. any) comparison did not yield significant results and the fact, that this was an exploratory study design. Nevertheless, this finding warrants further investigation.

As a positive takeaway the fact that happiness is by far the most common response in our survey is reassuring und clearly points to the resilience of this age group.

There are several limitations to our study. The sample size of around 180 infected individuals is not large enough to detect rare symptoms and a screening questionnaire cannot reliably compare the severity of symptoms in affected individuals. Furthermore, our questionnaire concentrated on neurocognitive, general pain and mood symptoms. Symptoms like a persistent sore throat, persistent cough or chest tightness and an altered sense of smell/ taste were not included.

However, our survey covers a variety of symptoms reported in the context of Long-COVID19 and having a control group of age-matched peers who never had a SARS-CoV-2 infection adds valuable information to the Long-COVID19 discussion that is urgently needed.

## Conclusion

In our cohort of adolescents more than one third reported the presence of at least one neurocognitive, pain or mood symptom with tenseness, listlessness and difficulties concentrating being reported most commonly. However, there was no statistical difference comparing the reported symptoms between seropositive students -with mild to asymptomatic courses of SARS-CoV-2 infections -and seronegative students. Leading to the conclusion that symptoms of Long-COVID19 might be less common than previously assumed and emphasizing on the impact of pandemic-associated symptoms regarding the well-being and mental health of young adolescents.

## Research in context

### Evidence before this study

We searched PubMed for articles published between January 1, 2020, and May 1, 2021, using the search terms (“Long-Covid19”) AND (“adolescent”) AND (“children”). We identified 1 relevant cross sectional study and 1 case series. Persisting symptoms up to 120 days after the SARS-CoV-2 infection were reported in at least 50% of children and adolescents.

### Added value of this study

By adding a control group this study documents that there is no significant difference in the prevalence of neurocognitive, pain and mood symptoms in seropositive compared to seronegative adolescents. This suggests that pandemic-and lockdown-associated factors affect the mental health of adolescents more than infection-associated factors.

### Implications of all the available evidence

These findings add relevant new data that will help to inform scientists, public health authorities and policy makers in regard to future policy measures in an ongoing pandemic.

## Data Availability

Deidentified individual participant data will be made available, in addition to study protocols, the statistical analysis plan, and the informed consent form. The data will be made available upon publication to researchers who provide a methodologically sound proposal for use in achieving the goals of the approved proposal. Proposals should be submitted to corresponding author.

## Acknowledgements

We thank the Federal State of Saxony for supporting this study by a financial grant.

We thank J. Schneider for her support and excellent organization of the study visit March/ April 2021.

We thank J. Herrmann and K. Jackisch for their great support in collecting all samples.

## Contributors

J.A and R.B. designed the study and wrote the protocol. J.A., J.B., M.W., J.R. and R.B. designed the Long-COVID19 Survey. J.A., J.B., C.K., L.H., E.K., L.S., P.C. collected samples. J.A., J.B. and M.W. analyzed and verified the data. J.A., J.B. and M.W. wrote the manuscript. M.W., R.B., C.K., L.H., E.K., L.S., P.C. reviewed the manuscript.

All corresponding authors had full access to all the data in the study and had final responsibility for the decision to submit for publication.

## Abbreviations

COVID-1: Coronavirus disease 2019
ELISA: enzyme-linked immunosorbent assay
IgG: Immunoglobulin G
IQR: Interquartile Range
PCR: Polymerase Chain Reaction
SARS-CoV-2: severe acute respiratory syndrome coronavirus type 2

---

**Supplementals - Figure 1).**
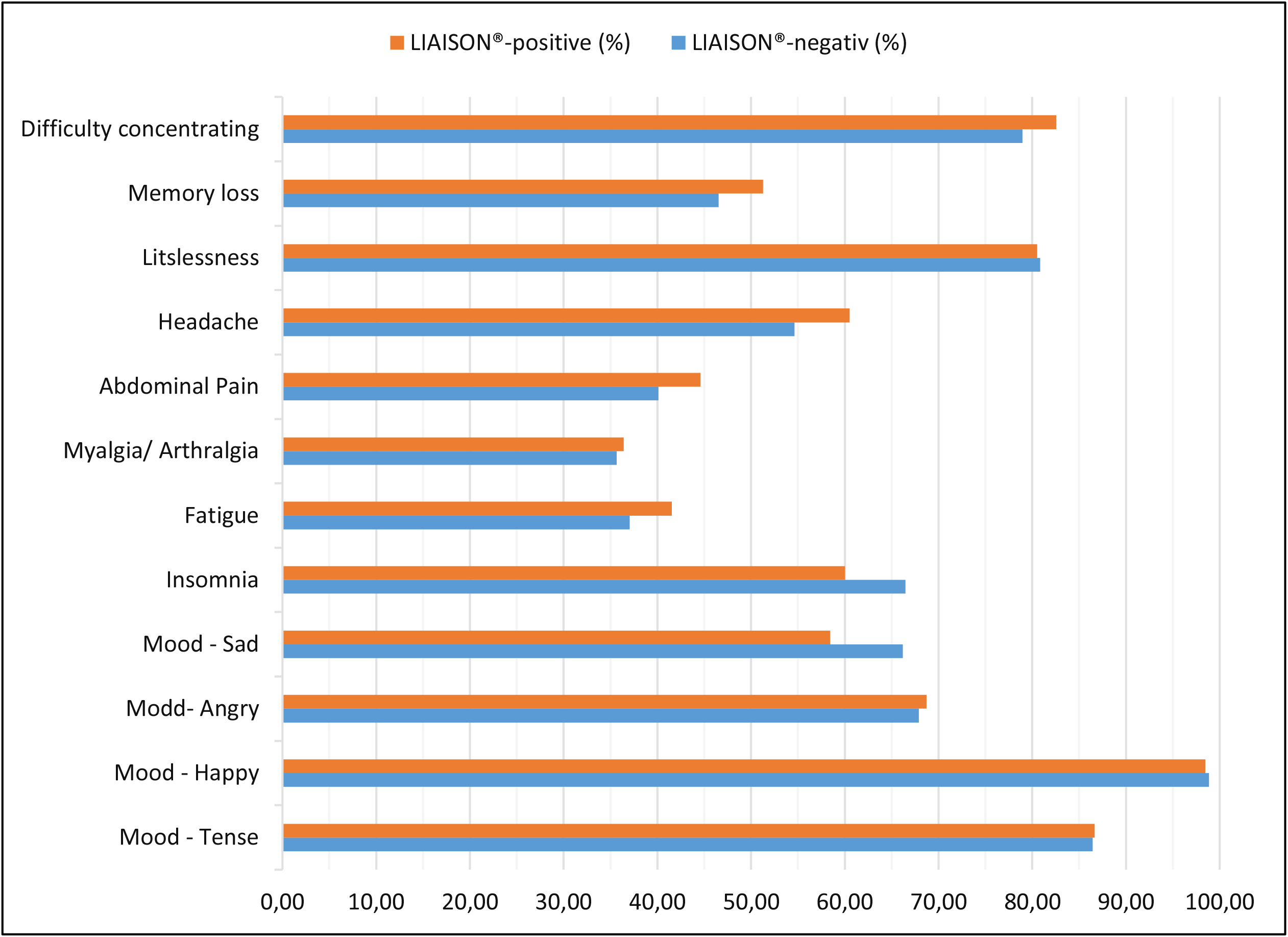
*Prevalence of neurocognitive, pain and mood symptoms in LIAISON®-negativ and positive study participants (Fisher’s exact test:* n = 1553, * significant at level 0·05)

